# Latent Profiles of Stigma and Their Influencing Factors in Post-Mastectomy Breast Cancer Patients: A Conservation of Resources Theory Perspective

**DOI:** 10.64898/2026.08.04.26359662

**Authors:** Lin Xia, Lin Tan, Shouqi Zheng, Ying Han, Lingyi Xiao, Jijun Wu, Lin He

## Abstract

**Objective:** To identify the latent profiles of stigma among breast cancer patients following modified radical mastectomy and to analyze their influencing factors, thereby providing an empirical foundation for tailored psychological nursing interventions.

**Methods:** A convenience sampling strategy was employed to recruit 276 post-mastectomy breast cancer patients from the breast surgery department of a tertiary care hospital in Sichuan Province between January 2026 and May 2026. Data were collected using a general demographic questionnaire, the Social Impact Scale (SIS), the Self-Compassion Scale (SCS), and the Multidimensional Scale of Perceived Social Support (PSSS). Item scores of the SIS served as explicit indicators for latent profile analysis (LPA). Univariate analysis and multinomial logistic regression were conducted to identify independent predictors associated with stigma profile membership.

**Results:** The total SIS score among post-mastectomy breast cancer patients was 53.88±7.90, indicating a moderate level of stigma. LPA yielded three distinct latent profiles: the low-stigma profile (22.1%), moderate-stigma profile (57.2%), and high-stigma profile (20.7%). Multinomial logistic regression analysis demonstrated that age, educational level, employment status, monthly per capita income, affected upper-limb paresthesia, clinical TNM stage, self-compassion, and perceived social support were independent predictors of stigma profile membership (all *P*<0.05).

**Conclusion:** Stigma among post-mastectomy breast cancer patients exhibits substantial latent heterogeneity, categorizable into three distinct psychological profiles. Healthcare professionals should pay particular attention to patients who are younger, more highly educated, face heavier economic burdens, present with affected upper-limb paresthesia, or are diagnosed with advanced TNM stages. Implementing stratified and individual-tailored interventions focused on cultivating self-compassion and bolstering social support is essential to attenuate stigma and foster postoperative psychological recovery.

## Introduction

According to the International Agency for Research on Cancer, breast cancer remains one of the most common malignancies and a leading cause of cancer-related mortality among women worldwide, with approximately 2.43 million new cases and 694,000 deaths reported in 2024[1]. Asia accounts for over 40% of the global breast cancer burden[1], with China recording around 375,000 new cases, ranking highest in the region[2]. Advances in early screening technologies and multimodal treatment strategies have significantly improved the 5-year survival rate of breast cancer patients to 83.2%[3, 4]. Consequently, clinical management strategies are progressively shifting from merely extending survival to optimizing long-term quality of life and psychosocial adaptation.

Modified radical mastectomy remains one of the primary surgical interventions for breast cancer[5]. However, because the breast is a significant secondary sexual characteristic in women, its surgical removal results in irreversible physical alterations and profound disruptions to body image and female identity[6]. Postoperatively, patients frequently experience adverse psychological distress, including anxiety, depression, reduced self-esteem, and social withdrawal[7, 8], which further impairs their functional recovery and quality of life. Among these challenges, stigma—a key psychosocial stressor—has emerged as a major barrier to long-term rehabilitation. Disease-related stigma refers to the psychological experience of social rejection, negative evaluation, and internalized shame associated with a condition, encompassing dimensions such as social rejection, social isolation, financial insecurity, and internalized shame[9]. Emerging evidence indicates that heightened stigma is strongly associated with severe depression, anxiety, fear of cancer recurrence, non-adherence to treatment, and diminished quality of life[7]. Persistent stigma not only erodes patients’ sense of self-worth and adaptive capacity but also impedes the restoration of social functioning, ultimately hindering long-term recovery following surgery.

According to Conservation of Resources (COR) theory, individuals strive to obtain, retain, and protect their valued resources, including time, energy, and emotional well-being; psychological stress and maladaptive outcomes occur when these resources are threatened, lost, or fail to yield expected returns following investment[10]. For patients undergoing modified radical mastectomy for breast cancer, surgical loss of the breast, physical functional limitations, and altered social roles can lead to a continuous depletion of vital resources, such as bodily integrity, self-esteem, female identity, and interpersonal relationships, thereby triggering a cascade of psychological stress responses. From the perspective of COR theory, disease-related stigma can be conceptualized as a negative psychological manifestation resulting from cumulative resource loss. The theory further posits that resources typically exist in "resource caravans," where abundant resource reservoirs facilitate the acquisition of additional resources, creating a gain spiral, whereas resource deficits predispose individuals to continuous loss spirals[11]. Consequently, an individual’s social support and psychological traits represent a dynamic balance between external resource acquisition and internal resource accumulation. Self-compassion—defined as an attitude of understanding, acceptance, and kindness toward oneself when facing flaws, failures, or suffering[12]—functions as a crucial internal personal resource. Higher levels of self-compassion mitigate self-criticism and shame cognitions, promote emotional recovery, and facilitate trauma acceptance, thereby buffering against illness-induced resource depletion[13]. Conversely, perceived social support reflects an individual’s subjective perception of support from family, friends, and other social connections, serving as a key external social resource[14]. Adequate social support not only provides emotional solace and practical assistance but also strengthens patients’ confidence in coping with disease and alleviates illness-related stress[15]. Based on these premises, we hypothesize that self-compassion and perceived social support exert an interactive effect on stigma; specifically, these internal and external resources may operate synergistically to further reduce stigma beyond their independent protective effects. However, this potential interactive mechanism has not yet been empirically validated in post-mastectomy breast cancer patients.

Current research on stigma among patients with breast cancer predominantly relies on variable-centered approaches, such as correlation and regression analyses, which tend to overlook potential subgroups and heterogeneity within patient populations. In contrast, latent profile analysis (LPA) is an person-centered statistical technique that models relationships among continuous observed variables through a latent categorical variable, thereby identifying distinct homogeneous subgroups to facilitate tailored cross-group comparative analyses and inform precision nursing strategies[16]. Although preliminary evidence has confirmed the heterogeneity of stigma in breast cancer patients[17], previous studies have primarily examined single-variable mechanisms—such as the mediating role of self-efficacy between stigma and sleep quality—without systematically investigating how the "synergistic allocation of internal and external resources" predicts distinct stigma profiles within a comprehensive theoretical framework. In summary, despite the emphasis of COR theory on the synergistic effects of resource caravans, empirical evidence remains scarce regarding how internal personal resources (self-compassion) and external social resources (perceived social support) jointly shape latent stigma profiles in patients following modified radical mastectomy. To address this gap, guided by COR theory, this study employed LPA to identify potential categories of stigma among post-mastectomy breast cancer patients and further examined the predictive effects of self-compassion, perceived social support, and relevant demographic and clinical characteristics on subgroup membership. By delineating these associations, this study aims to provide healthcare professionals with objective criteria for early identification of high-risk populations, thereby offering a solid theoretical foundation and empirical evidence to design tailored nursing interventions that integrate psychological adjustment with social support reinforcement.

## Method

### Participants and sample size

A convenience sampling method was used to recruit patients with breast cancer who underwent modified radical mastectomy at the breast surgery department of a Class III Grade A hospital in Sichuan Province between January 2026 and May 2026. The inclusion criteria were as follows: (1) Histologically/pathologically confirmed breast cancer[18];(2) Voluntary participation with the capability to independently complete the questionnaire; (3) Age≥18 years; (4) History of modified radical mastectomy; (5) Clinical TNM stage I-III.The exclusion criteria were: (1) Co-occurrence of other primary malignancies; (2) Presence of psychiatric disorders or severe systemic comorbidities that precluded effective communication and survey completion. Regarding sample size considerations for latent profile analysis (LPA), previous simulation studies have demonstrated that a sample size of N≥250 yields stable, robust classification parameters and reliable model fit indices[19]. Furthermore, general guidelines for multinomial logistic regression recommend a minimum of 10 participants per candidate predictor variable[20]. Given that 17 independent variables were included in the multivariable analysis, a minimum of 170 participants was required to maintain sufficient statistical power. The final sample size of 276 participants in this study satisfied the analytical requirements for both LPA and multivariable modeling. The study protocol was approved by the Ethics Review Board of Deyang City People’s Hospital (No. 2026-04-050-K01, dated January 25th, 2026).

### Measures

#### Sociodemographic and clinical characteristics

A researcher-developed questionnaire was used to collect participants’ demographic and clinical characteristics. Demographic information included age, place of residence, educational level, marital status, employment status, monthly per capita income, medical payment method, smoking history, and alcohol consumption history. Clinical information included comorbidities, postoperative complications, molecular subtype, disease duration, and TNM stage.

#### Social impact scale (SIS)

The scale was developed by Fife et al.[9] and translated into Chinese by Pan et al.[21], has been widely used to assess stigma among patients with cancer. The scale comprises four dimensions: financial insecurity (3 items), social rejection (9 items), internalized shame (5 items), and social isolation (7 items), with a total of 24 items. Items are rated on a 4-point Likert scale ranging from 1 (strongly disagree) to 4 (strongly agree), with total scores ranging from 24 to 96. Higher scores indicate greater perceived stigma. Total scores of ≤39, 40-59, and ≥60 indicate low, moderate, and high levels of stigma, respectively. Previous studies have demonstrated satisfactory psychometric properties of the SIS, with Cronbach’s α coefficients ranging from 0.85-0.90. In the present study, the Cronbach’s α coefficient was 0.850.

#### Self-compassion scale (SCS)

The scale was originally developed by Neff in 2003[12] and subsequently translated and culturally adapted into Chinese by Chen et al.[22] The Chinese version comprises 26 items across six dimensions: self-kindness (5 items), common humanity (4 items), mindfulness (4 items), self-judgment (5 items), isolation (4 items), and over-identification (4 items). The dimensions of self-judgment, isolation, and over-identification are reverse scored. Items are rated on a 5-point Likert scale ranging from 1 (almost never) to 5 (almost always), with total scores ranging from 26 to 130. Higher scores indicate higher levels of self-compassion. In the present study, the Cronbach’s α coefficient was 0.915.

#### Perceived social support scale (PSSS)

The scale was originally developed by Zimet et al.[23] in 1987 and subsequently translated and culturally adapted into Chinese by Jiang.[24] The scale consists of 12 items across three dimensions: family support (4 items), friend support (4 items), and support from significant others (4 items). Items are rated on a 7-point Likert scale ranging from 1 (very strongly disagree) to 7 (very strongly agree), with total scores ranging from 12 to 84. Higher scores indicate greater perceived social support. Total scores can be categorized as low (12-36), moderate (37-60), and high (61-84) levels of perceived social support. In the present study, the Cronbach’s α coefficient was 0.872.

### Data collection

To ensure consistency in data collection, all survey coordinators underwent standardized, offline pre-study training covering the study objectives, item interpretations, informed consent protocols, and contingency handling procedures. Survey coordinators presented a QR code linked to an online survey platform (Wenjuanxing) to eligible post-mastectomy breast cancer patients. Upon scanning the QR code with smartphones, participants completed the electronic questionnaire, which enforced mandatory responses for all items and restricted submissions to a single response per IP address and device to prevent duplicate entries. To accommodate older adults and individuals unfamiliar with smart devices, a parallel face-to-face paper-based survey was conducted. Trained research nurses distributed paper questionnaires in inpatient wards or outpatient waiting areas, providing on-site instructions. Patients completed the questionnaires independently, which were immediately retrieved upon completion. For patients unable to complete the survey independently due to visual impairment, upper limb dysfunction, or limited literacy, research nurses read each item aloud using a neutral tone and recorded the responses verbatim, strictly avoiding leading explanations to ensure response objectivity. The survey instruments comprised demographic characteristics, disease-related variables, the revised SIS, PSSS, and SCS. Disease-related clinical data were extracted directly from the Hospital Information System (HIS) by the research team. Throughout the data collection period, coordinators were present to address any item-clarification queries. Following retrieval, all questionnaires underwent dual data verification by the research team, and entries with excessively short completion times, repetitive response patterns, or incomplete data were excluded. Ultimately, 290 questionnaires were distributed, and 276 valid responses were recovered, yielding an effective response rate of 95.17%.

### Statistical analysis

Data were independently entered by two researchers and cross-checked to ensure accuracy. Statistical analyses were performed using SPSS (version 26.0; IBM Corp., Armonk, NY, USA) and Mplus (version 8.3; Muthén & Muthén, Los Angeles, CA, USA). Continuous variables adhering to a normal distribution were presented as means±standard deviations (X±S), whereas non-normally distributed continuous variables were expressed as medians and interquartile ranges [M(P25, P75)]. Categorical variables were reported as absolute frequencies and percentages (%), with inter-group comparisons evaluated using the χ^2^ test or Fisher’s exact test, as appropriate. Correlations between continuous variables were assessed using Pearson or Spearman rank correlation analysis.Latent profile analysis (LPA) was conducted using Mplus 8.3 to identify potential subgroups of stigma among post-mastectomy breast cancer patients. Model fit was evaluated using several statistical indicators: Akaike Information Criterion(AIC), Bayesian Information Criterion(BIC), Sample-Size-Adjusted BIC(aBIC), Entropy, Lo-Mendell-Rubin Likelihood Ratio Test(LMRT), and Bootstrap Likelihood Ratio Test(BLRT). Lower AIC, BIC, and aBIC values indicate superior model fit. Entropy values range from 0 to 1, with higher values indicating greater classification precision. Statistically significant *P*-values for LMRT and BLRT suggest that the k-class model provides a significantly better fit than the (k-1)-class model.To explore the factors associated with distinct stigma profile memberships, multinomial logistic regression analysis was executed. Prior to model construction, to minimize potential multicollinearity between main effects and their interaction term, the continuous independent variables—total SCS score and total PSSS score—were mean-centered (c_SCS and c_PSSS). The interaction product term (c_SCS * c_PSSS) was subsequently calculated and incorporated into the regression model. Statistical significance was set at a two-tailed α = 0.05.

## Results

### Scores of the SIS, SCS, and PSSS among patients following modified radical mastectomy for breast cancer

The mean total SIS score among patients following modified radical mastectomy for breast cancer was (53.88±7.90), corresponding to a mean item score of (2.25±0.33) and indicating a moderate level of stigma. Among the four dimensions, internalized shame had the highest mean item score (2.49±0.45), followed by social isolation (2.37±0.41) and financial insecurity (2.35±0.53), whereas social rejection showed the lowest score (1.98±0.38). The demographic and clinical characteristics of the participants are presented in Table 1, and correlations among stigma, self-compassion, and perceived social support are shown in Table 2.

**Table 1.** Demographic and clinical characteristics and univariate analysis of latent stigma profiles among patients following modified radical mastectomy for breast cancer

| Variables | Low stigma profile<br>(n=61) | Moderate stigma profile<br>(n=158) | High stigma profile<br>(n=57) | Statistic | P value |
| --- | --- | --- | --- | --- | --- |
| Age(years) |  |  |  |  |  |
| <50 | 19(31.1) | 91(57.6) | 47(82.5) | 35.942<sup>a</sup> | <0.001 |
| 50-60 | 27(44.3) | 53(33.5) | 8(14.0) |  |  |
| >60 | 15(24.6) | 14(8.9) | 2(3.5) |  |  |
| Place of residence |  |  |  |  |  |
| Urban area | 31(50.8) | 85(53.8) | 24(42.1) | 4.064<sup>a</sup> | 0.397 |
| Rural area | 21(34.4) | 46(29.1) | 18(31.6) |  |  |
| Township/County area | 9(14.8) | 27(17.1) | 15(26.3) |  |  |
| Educational level |  |  |  |  |  |
| High school or above | 10(16.4) | 72(45.6) | 47(82.5) | 51.862<sup>a</sup> | <0.001 |
| Junior high school or below | 51(83.6) | 86(54.4) | 10(17.5) |  |  |
| Marital status |  |  |  |  |  |
| Single | 1(1.6) | 4(2.5) | 0(0.0) | 3.202<sup>b</sup> | 0.804 |
| Married | 55(90.2) | 143(90.5) | 53(93.0) |  |  |
| Divorced or separated | 2(3.3) | 8(5.1) | 2(3.5) |  |  |
| Widowed | 3(4.9) | 3(1.9) | 2(3.5) |  |  |
| Living arrangement |  |  |  |  |  |
| Living with spouse | 27(45.0) | 40(25.8) | 19(33.9) | 12.553<sup>a</sup> | 0.51 |
| Living with spouse and children | 21(35.0) | 76(49.0) | 18(32.1) |  |  |
| Living alone | 3(5.0) | 8(5.2) | 2(3.6) |  |  |
| Others | 9(15.0) | 31(20.0) | 17(30.4) |  |  |
| Employment status |  |  |  |  |  |
| Employed | 25(41.0) | 38(24.1) | 2(3.5) | 33.159<sup>a</sup> | <0.001 |
| Self-employed | 19(31.1) | 50(31.6) | 22(38.6) |  |  |
| Retired | 12(19.7) | 48(30.4) | 14(24.6) |  |  |
| Farmer | 5(8.2) | 22(13.9) | 19(33.3) |  |  |
| Monthly per capita income (RMB) |  |  |  |  |  |
| <3000 | 19(31.1) | 79(50.0) | 48(84.2) | 36.975<sup>b</sup> | <0.001 |
| 3000-5000 | 36(59.0) | 72(45.6) | 8(14.0) |  |  |
| >5000 | 6(9.8) | 7(4.4) | 1(1.8) |  |  |
| Health insurance type |  |  |  |  |  |
| Urban Employee Basic Medical Insurance | 36(59.0) | 92(58.2) | 32(56.1) | 3.596<sup>b</sup> | 0.729 |
| Urban-Rural Resident Basic Medical Insurance | 19(31.1) | 39(24.7) | 18(31.6) |  |  |
| Commercial insurance | 0(0.0) | 4(2.5) | 0(0.0) |  |  |
| Self-pay | 6(9.8) | 23(14.6) | 7(12.3) |  |  |
| Comorbidities |  |  |  |  |  |
| No | 47(77.0) | 140(88.6) | 46(80.7) | 5.225<sup>a</sup> | 0.073 |
| Yes | 14(23.0) | 18(11.4) | 11(19.3) |  |  |
| Sensory abnormalities in the affected upper limb |  |  |  |  |  |
| No | 54(88.5) | 91(57.6) | 17(29.8) | 42.064<sup>a</sup> | <0.001 |
| Yes | 7(11.5) | 67(42.4) | 40(70.2) |  |  |
| Perceived disease severity |  |  |  |  |  |
| Severe | 24(39.3) | 52(32.9) | 31(54.4) | 8.148<sup>a</sup> | 0.017 |
| Non-severe | 37(60.7) | 106(67.1) | 26(45.6) |  |  |
| Smoking history |  |  |  |  |  |
| No | 60(98.4) | 150(94.9) | 53(93.0) | 1.960<sup>b</sup> | 0.364 |
| Yes | 1(1.6) | 8(5.1) | 4(7.0) |  |  |
| Alcohol consumption history |  |  |  |  |  |
| No | 58(95.1) | 141(89.2) | 48(84.2) | 3.729<sup>a</sup> | 0.155 |
| Yes | 3(4.9) | 17(10.8) | 9(15.8) |  |  |
| Time since diagnosis (years) |  |  |  |  |  |
| <1 | 26(42.6) | 55(34.8) | 23(40.4) | 3.164<sup>a</sup> | 0.531 |
| 1-5 | 23(37.7) | 77(48.7) | 22(38.6) |  |  |
| >5 | 12(19.7) | 26(16.5) | 12(21.1) |  |  |
| TNM stage |  |  |  |  |  |
| I | 23(37.7) | 19(12.0) | 2(3.5) | 55.506<sup>a</sup> | <0.001 |
| II | 32(52.5) | 75(47.5) | 16(28.1) |  |  |
| III | 6(9.8) | 64(40.5) | 39(68.4) |  |  |
| Total SCS score | 96.00(90.50,103.00) | 92.00(84.00,99.25) | 83.00(76.50,91.00) | 39.328<sup>c</sup> | <0.001 |
| Total PSSS score | 61.00(57.00,67.00) | 58.00(53.00,63.00) | 52.00(46.00,58.00) | 35.517<sup>c</sup> | <0.001 |

<sup>a</sup> $\chi^2$ = chi square; <sup>b</sup>Fisher's exact test; <sup>c</sup>H=Kruskal-Wallis test; SCS=self-compassion scale;
PSSS=perceived social support scale

**Table 2.** Correlation Among SIS, SCS, and PSSS Scores in Postoperative Breast Cancer Patients

|  | Total SIS score | Financial<br>insecurity | Social rejection | Internalized shame | Social isolation |
| --- | --- | --- | --- | --- | --- |
| Total SCS score | -0.292<sup>①</sup> | -0.296<sup>①</sup> | -0.254<sup>①</sup> | -0.163<sup>①</sup> | -0.222<sup>①</sup> |
| Self-judgment | 0.211<sup>①</sup> | 0.226<sup>①</sup> | 0.131<sup>①</sup> | 0.143<sup>①</sup> | 0.199<sup>①</sup> |
| Over-identification | 0.260<sup>①</sup> | 0.241<sup>①</sup> | 0.224<sup>①</sup> | 0.153<sup>①</sup> | 0.202<sup>①</sup> |
| Common humanity | -0.256<sup>①</sup> | -0.179<sup>①</sup> | -0.239<sup>①</sup> | -0.188<sup>①</sup> | -0.165<sup>①</sup> |
| Isolation | 0.233<sup>①</sup> | 0.187<sup>①</sup> | 0.210<sup>①</sup> | 0.140<sup>①</sup> | 0.188<sup>①</sup> |
| Self-kindness | -0.169<sup>①</sup> | -0.254<sup>①</sup> | -0.148<sup>①</sup> | -0.061 | -0.11 |
| Mindfulness | -0.113 | -0.170<sup>①</sup> | -0.134<sup>①</sup> | -0.001 | -0.069 |
| Total PSSS score | -0.321<sup>①</sup> | -0.226<sup>①</sup> | -0.266<sup>①</sup> | -0.204<sup>①</sup> | -0.299<sup>①</sup> |
| Family support | -0.234<sup>①</sup> | -0.177<sup>①</sup> | -0.209<sup>①</sup> | -0.115 | -0.231<sup>①</sup> |
| Friend support | -0.216<sup>①</sup> | -0.173<sup>①</sup> | -0.155<sup>①</sup> | -0.176<sup>①</sup> | -0.176<sup>①</sup> |
| Other Support | -0.297<sup>①</sup> | -0.181<sup>①</sup> | -0.261<sup>①</sup> | -0.175<sup>①</sup> | -0.295<sup>①</sup> |

<sup>①</sup> $P < 0.05$ 。

### Common method bias test

Harman’s single-factor test via exploratory factor analysis was conducted to assess potential common method bias. The results identified 16 factors with eigenvalues≥1. The first unrotated factor accounted for 16.85% of the total variance, which is substantially below the critical threshold of 40%. These findings indicate that common method bias was not a serious threat to the validity of the data in this study.

### Latent profiles and naming of stigma in post-mastectomy breast cancer patients

Item scores from the 24-item SIS served as explicit indicators for exploratory LPA, sequentially testing 1- to 5-profile models. Fit indices for all candidate models are summarized in Table 3. As the number of profiles increased, the AIC and BIC values steadily decreased, while Entropy values across all models consistently exceeded 0.90, indicating high classification precision. Model 3 exhibited lower AIC, BIC, and sample-size-aBIC values compared with lower-order models, alongside statistically significant LMRT and BLRT *P*-values (*P*<0.05). Although Model 4 had slightly lower AIC/BIC, the LMRT test was not significant (*P*=0.368), indicating the 3-profile solution is statistically more parsimonious. Balancing model parsimony and theoretical interpretability, Model 3 was identified as the optimal latent profile solution. The mean item score patterns for Model 3 are illustrated in Fig 1, based on which the three distinct subgroups were defined. Profile 1 (n=61, 22.1%) scored the lowest across all SIS dimensions, reflecting robust self-efficacy and sufficient perceived external support; this subgroup was named the "low-stigma profile". Profile 2 (n=158, 57.2%) presented moderate scores across all dimensions, with internalized shame being the most prominent feature. Because its overall psychological state fell between the low and high groups, it was labeled the "moderate-stigma profile". Profile 3 (n=57, 20.7%) exhibited the highest scores across all dimensions, indicating pervasive psychosocial functional impairment, and was designated the "high-stigma profile". Total SIS scores and subscale scores differed significantly among the three profiles (*P*<0.001; Table 4), confirming substantial latent heterogeneity in stigma among post-mastectomy breast cancer patients.

**Figure 1.**
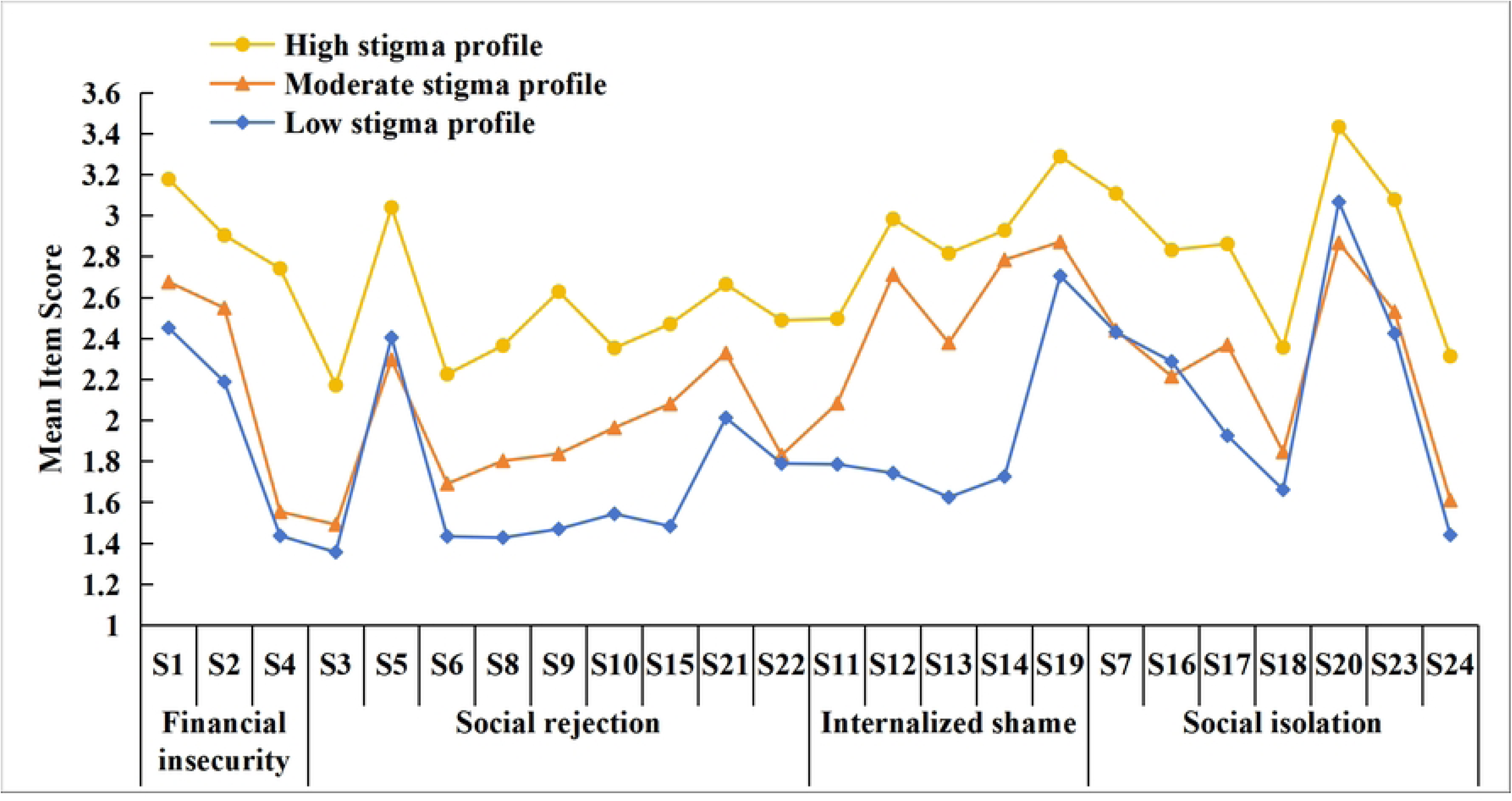
Characteristics of Latent Profiles of Stigma Among Postoperative Breast Cancer Patients

**Table 3.**
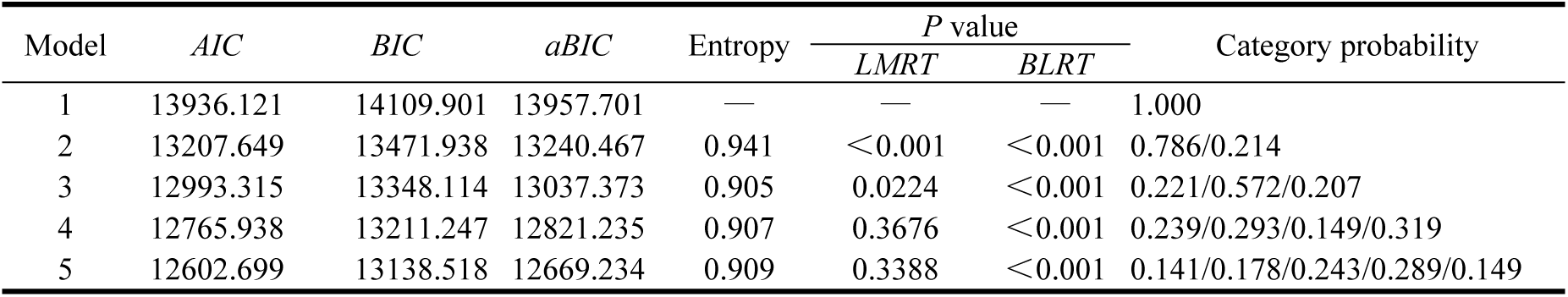
Fit Indices for Latent Profile Models of Stigma in Postoperative Breast Cancer Patients

**Table 4.** Comparison of SIS, SCS, and PSSS Scores Across Latent Stigma Profiles Among Postoperative Breast Cancer Patients

| Profile | *N* | Total SIS score |  | Financial insecurity |  | Social rejection |  | Internalized shame |  | Social isolation |  |
| --- | --- | --- | --- | --- | --- | --- | --- | --- | --- | --- | --- |
|  |  | score | Mean Item | score | Mean Item | score | Mean Item | score | Mean Item | score | Mean Item |
|  |  |  | Score |  | Score |  | Score |  | Score |  | Score |
| 1 | 61 | 45.72±0.46 | 1.91±0.02 | 6.07±0.19 | 2.02±0.06 | 14.89±0.23 | 1.65±0.03 | 15.25±0.30 | 1.91±0.03 | 9.52±0.14 | 2.18±0.04 |
| 2 | 158 | 52.75±0.33 | 2.20±0.01 | 6.77±0.10 | 2.26±0.03 | 17.30±0.19 | 1.92±0.02 | 15.87±0.18 | 2.56±0.02 | 12.81±0.11 | 2.27±0.03 |
| 3 | 57 | 65.75±0.65 | 2.74±0.03 | 8.82±0.15 | 2.94±0.05 | 22.42±0.38 | 2.49±0.04 | 19.98±0.29 | 2.91±0.05 | 14.53±0.25 | 2.85±0.04 |
| *H* value |  | 184.051 |  | 93.655 |  | 144.599 |  | 97.829 |  | 144.797 |  |
| *P* value |  | <0.001 |  | <0.001 |  | <0.001 |  | <0.001 |  | <0.001 |  |

Note: Because the scores of certain dimensions in 1–3 exhibited heterogeneity of variance across the three profiles, the Kruskal-Wallis *H* test was uniformly applied.

### Univariate analysis of stigma latent profiles

Univariate analysis demonstrated that the distribution of stigma latent profiles among post-mastectomy breast cancer patients varied significantly by age, educational level, employment status, monthly per capita income, affected upper-limb paresthesia, perceived disease severity, clinical TNM stage, self-compassion, and perceived social support (all *P*<0.05; Table 1).

### Multivariable analysis of stigma latent profiles

Multinomial logistic regression analysis was performed using the three latent profiles of stigma as the dependent variable (with the low-stigma profile serving as the reference group). Independent variables included demographic and clinical factors that showed statistical significance in univariate analysis, alongside mean-centered self-compassion (c_SCS), mean-centered perceived social support (c_PSSS), and their interaction term (c_SCS * c_PSSS). Variable assignment and coding schemes are detailed in Table 5. Multivariable results revealed that age, educational level, employment status, monthly per capita income,affected upper-limb paresthesia, clinical TNM stage, self-compassion, and perceived social support were independent predictors of stigma profile membership (all *P*<0.05; Table 6). Notably, the interaction term between self-compassion and perceived social support (c_SCS*c_PSSS) was not statistically significant across profile comparisons (*P*>0.05), indicating that internal psychological resources and external social resources operate independently rather than synergistically in predicting stigma profile membership.

**Table 5.** Coding scheme for independent variables

| Variable | Value assignment |
| --- | --- |
| Age(years) | $< 50 = (0,0)$; $50 \sim 60 = (1,0)$; $> 60 = (0,1)$ |
| Educational level | High school or above=0; Junior high school or below=1 |
| Employment status | Employed=(0,0,0); Self-employed=(1,0,0); Retired=(0,1,0); Farmer=(0,0,1) |
| Monthly per capita income (RMB) | $< 3000 = (0,0)$; $3000 \sim 5000 = (1,0)$; $> 5000 = (0,1)$ |
| Sensory abnormalities in the affected upper limb | Yes=0; No=1 |
| Perceived disease severity | Severe=0; Non-severe=1 |
| TNM stage | I=(0,0); II=(1,0); III=(0,1) |
| SCS | $c\_SCS = SCS - \overline{SCS}$ |
| PSSS | $c\_PSSS = PSSS - \overline{PSSS}$ |
| SCS*PSSS | $c\_SCS * c\_PSSS$ |

**Table 6.** Factors Associated With Latent Stigma Profile Membership Among Postoperative Breast Cancer Patients: A Multinomial Logistic Regression Analysis

| Variables | Moderate stigma profile vs.Low stigma profile |  |  |  |  |  | High stigma profile vs.Low stigma profile |  |  |  |  |  |
| --- | --- | --- | --- | --- | --- | --- | --- | --- | --- | --- | --- | --- |
| | $\beta$ | SE | Wald $\chi^2$ | P | OR | 95%CI | $\beta$ | SE | Wald $\chi^2$ | P | OR | 95%CI |
| Age(years) |  |  |  |  |  |  |  |  |  |  |  |  |
| 50-60 | -0.398 | 0.447 | 0.794 | 0.373 | 0.672 | (0.280, 1.612) | -1.112 | 0.699 | 2.527 | 0.112 | 0.329 | (0.084, 1.295) |
| >60 | -1.475 | 0.609 | 5.859 | 0.015 | 0.229 | (0.069, 0.755) | -3.154 | 1.192 | 6.996 | 0.008 | 0.043 | (0.004, 0.442) |
| Educational level |  |  |  |  |  |  |  |  |  |  |  |  |
| High school or above | 1.071 | 0.469 | 5.214 | 0.022 | 2.918 | (1.164, 7.317) | 2.362 | 0.654 | 13.054 | <0.001 | 10.617 | (2.947, 38.244) |
| Employment status |  |  |  |  |  |  |  |  |  |  |  |  |
| Self-employed | 0.651 | 0.479 | 1.845 | 0.174 | 1.917 | (0.750, 4.903) | 2.847 | 1.054 | 7.288 | 0.007 | 17.231 | (2.181, 136.111) |
| Retired | 0.338 | 0.548 | 0.379 | 0.538 | 1.401 | (0.478, 4.105) | 1.638 | 1.101 | 2.214 | 0.137 | 5.146 | (0.595, 44.529) |
| Farmer | -0.495 | 0.722 | 0.470 | 0.493 | 0.609 | (0.148, 2.511) | 1.494 | 1.189 | 1.579 | 0.209 | 4.456 | (0.433, 45.811) |
| Monthly per capita income (RMB) |  |  |  |  |  |  |  |  |  |  |  |  |
| 3000-5000 | -0.931 | 0.420 | 4.920 | 0.027 | 0.394 | (0.173, 0.897) | -2.190 | 0.648 | 11.439 | 0.001 | 0.112 | (0.031, 0.398) |
| >5000 | -1.015 | 0.912 | 1.239 | 0.266 | 0.362 | (0.061, 2.164) | -1.963 | 1.474 | 1.772 | 0.183 | 0.140 | (0.008, 2.527) |
| Sensory abnormalities in the affected upper limb | 2.141 | 0.530 | 16.297 | <0.001 | 8.506 | (3.008, 24.049) | 3.570 | 0.684 | 27.205 | <0.001 | 35.500 | (9.283, 135.757) |
| Perceived disease severity |  |  |  |  |  |  |  |  |  |  |  |  |
| Severe | -0.555 | 0.402 | 1.908 | 0.167 | 0.574 | (0.261, 1.262) | 0.338 | 0.581 | 0.339 | 0.561 | 1.403 | (0.449, 4.384) |
| TNM stage |  |  |  |  |  |  |  |  |  |  |  |  |
| Stage II | 0.710 | 0.472 | 2.267 | 0.132 | 2.034 | (0.807, 5.128) | 0.849 | 1.144 | 0.550 | 0.458 | 2.336 | (0.248, 22.013) |
| Stage III | 2.214 | 0.658 | 11.326 | 0.001 | 9.156 | (2.521, 33.251) | 3.128 | 1.230 | 6.469 | 0.011 | 22.825 | (2.050, 254.199) |
| SCS | -0.042 | 0.023 | 3.363 | 0.067 | 0.959 | (0.917, 1.003) | -0.135 | 0.034 | 15.646 | <0.001 | 0.874 | (0.817, 0.934) |
| PSSS | -0.090 | 0.030 | 8.821 | 0.003 | 0.914 | (0.862, 0.970) | -0.138 | 0.041 | 11.404 | 0.001 | 0.872 | (0.805, 0.944) |
| SCS*PSSS | 0.002 | 0.002 | 0.997 | 0.318 | 1.002 | (0.998, 1.007) | -0.001 | 0.003 | 0.108 | 0.743 | 0.999 | (0.992, 1.006) |

## Discussion

### Post-mastectomy stigma status and its latent heterogeneity

In the present study, the total stigma score among post-mastectomy breast cancer patients was 53.88±7.90, indicating a moderate level of stigma, which is consistent with previous findings[25].Among the subscales, the internalized shame dimension exhibited the highest item mean score (2.49±0.45), suggesting that stigma in this population primarily manifests as internalized self-deprecation rather than external rejection. This pattern may stem from the fact that surgical removal of the breast—a primary symbol of female identity—directly shatters the patient’s gender identity. Even when external appearances are restored via reconstructive surgery, eradicating the deep-seated internalized shame associated with bodily "incompleteness" remains challenging[26].

Latent profile analysis revealed three distinct subgroups, confirming significant heterogeneity in stigma among post-mastectomy breast cancer patients. The Moderate-Stigma Profile (Profile2, 57.2%): Representing the largest proportion of participants, this profile exhibited moderate scores across all dimensions, with internalized shame being the most prominent. Patients in this category are mostly in a transitional phase from acute postoperative stress to long-term adaptation; they tend to employ passive endurance and withdrawal strategies[27], lacking the capacity to actively recruit self-compassion or seek external social support. The Low-Stigma Profile (Profile1, 22.1%): Exhibiting the lowest scores across all dimensions, this profile reflects robust internal self-efficacy and sufficient external support perception. Notably, however, these patients displayed a relatively pronounced score in the social isolation domain. Rather than indicating maladaptation, this pattern may reflect a strategy of selective social withdrawal or defensive boundary construction during the early postoperative stage. By deliberately minimizing non-essential, high-risk social interactions, patients shield themselves from intrusive gaze and secondary psychological distress, thereby protecting their limited emotional resources from depletion. The High-Stigma Profile (Profile3, 20.7%): Demonstrating significantly higher scores across all dimensions compared to the other two profiles, this group manifested pervasive psychological and social functional impairment. Patients in this profile were predominantly younger, more highly educated, and presented with advanced TNM stages. They frequently exhibited marked social avoidance and relationship alienation, with some completely severing external social ties while simultaneously experiencing intense financial strain and role loss. From the perspective of Conservation of Resources (COR) theory, the observed latent heterogeneity reflects individual variation in the dynamic balance between resource loss and resource reservoirs. Patients in the high-stigma profile face a continuous loss spiral driven by physical disfigurement and social role disruption, whereas those in the low-stigma profile build an effective protective buffer supported by abundant internal and external resources. Consequently, healthcare professionals should implement stratified, precision interventions. Special emphasis should be placed on early psychological counseling and functional reconstruction for high-risk populations, alongside comprehensive psychosocial support to reduce social avoidance, alleviate self-deprecation, and promote holistic rehabilitation.

### Demographic factors associated with latent stigma profiles

#### Demographic factors

The multivariable analysis demonstrated that age was a significant predictor of stigma latent profile membership among post-mastectomy breast cancer patients. Specifically, patients aged >60 years were significantly more likely to be classified into the low-stigma profile (*OR*=0.229, *P*=0.015; *OR*=0.043, *P*=0.008). This finding may be attributed to the likelihood that older patients gradually develop higher acceptance of disease prognosis, face more relaxed social role expectations, and place reduced emphasis on alterations in post-surgical body image. Conversely, younger patients are typically in critical life stages involving career development, marital maintenance, and child-rearing[28]. They often maintain higher expectations regarding physical appearance and social image; consequently, the loss of abreast exerts a far more disruptive impact on their social roles and intimate relationships. Furthermore, younger individuals generally hold higher health expectations, rendering them more vulnerable to profound feelings of loss and anxiety when health is compromised, thereby intensifying disease-related stigma. In addition, our study revealed that patients with an educational level of high school or above were more likely to belong to the high-stigma profile (*OR*=2.918, *P*=0.022; *OR*=10.617, *P*<0.001), indicating heightened stigma among highly educated individuals, which aligns with previous findings[29]. From the perspective of Conservation of Resources (COR) theory, younger and more highly educated patients typically possess richer social expectations and career capital—conceptualized as condition resources. The physical disfigurement resulting from modified radical mastectomy inflicts a severe blow to their established social roles, triggering an acute stress response driven by anticipated resource loss. Moreover, although higher educational attainment facilitates active health information seeking, it also endows patients with a more thorough understanding of disease prognosis, recurrence risks, and potential complications, paradoxically exacerbating negative self-evaluations and excessive health anxieties. Accordingly, healthcare providers should pay tailored attention to the psychological well-being of younger and highly educated post-mastectomy patients. Clinicians should actively explore their emotional fluctuations and internal experiences. When delivering health education, clinicians should avoid overemphasizing disease severity or potential negative outcomes to prevent exacerbating psychological burdens. Instead, educational strategies should focus on fostering rational disease perceptions and promoting positive body acceptance, thereby guiding these patients toward a more objective and adaptive psychological transition.

Our study further revealed that self-employed patients were significantly more likely to belong to the high-stigma profile (*OR*=17.231, *P*=0.007). These individuals often lack stable income sources and comprehensive social security benefits. Combined with the ongoing medical expenses required during postoperative recovery, this financial strain exacerbates feelings of social alienation and insecurity triggered by the illness. Furthermore, self-employed individuals generally maintain relatively limited daily social networks and lack the structural social recognition and collegial support embedded in formal workplace environments[30]. Consequently, they are more prone to focusing on negative illness experiences, finding it difficult to gain psychological buffering through active social role participation. Regarding monthly per capita income, a moderate income level (3,000-5,000RMB) served as a protective factor against elevated stigma. Compared with patients earning <3,000RMB monthly, those in the moderate-income bracket exhibited significantly lower levels of stigma, aligning with the findings of Wang et al.[31]. Patients in lower-income categories experience significant economic burden and financial strain, which can substantially reduce their capacity to cope with disease-related psychological distress. These constraints restrict their access to rehabilitation resources, psychological support, and social engagement opportunities[32], thereby fostering a vicious cycle of "poverty induced by illness, and stigma exacerbated by poverty" that intensifies the stigma experience.Therefore, healthcare professionals should closely evaluate patients’ occupational and financial backgrounds. For self-employed and low-income patients, clinicians and medical social workers should integrate psychological counseling with practical assistance, such as facilitating linkages to medical assistance policies and charitable foundation resources, thereby mitigating the supplementary psychological burden imposed by financial distress.

#### Disease-related factors

Our findings revealed that clinical TNM stage III was an independent predictor of stigma profile membership among post-mastectomy breast cancer patients, with patients in stage III being significantly more likely to be classified into the moderate-or high-stigma profiles (*OR*=9.156, *P*<0.001; *OR*=22.825, *P*=0.011). This aligns with previous studies by Tan et al. and Yu et al.[15, 33]. A plausible explanation is that patients with advanced-stage breast cancer face more complex treatment decisions, elevated risks of recurrence and metastasis, greater financial burdens, and prolonged recovery trajectories. Simultaneously, treatment-related side effects—such as alopecia, weight fluctuations, and cancer-related fatigue—exacerbate negative self-evaluations of body image, while persistent uncertainty regarding prognosis weakens psychological resilience. These findings suggest that healthcare professionals should prioritize advanced-stage patients during routine perioperative and follow-up stigma screenings, implementing comprehensive strategies that combine symptom management, psychological support, and social reinforcement to foster adaptive disease perceptions and enhance long-term health outcomes.Additionally, this study incorporated affected upper-limb paresthesia as an independent predictor to explore the psychological impact of neuropathic somatic discomfort. Results demonstrated that upper-limb paresthesia was an independent predictor of stigma profile membership, substantially increasing the odds of belonging to the moderate- or high-stigma profiles (*OR*=8.506, *P*<0.001; *OR*=35.500, *P*<0.001). From the perspective of Conservation of Resources (COR) theory, somatic functional integrity serves as a foundational energy resource for maintaining daily psychological adaptation. Qualitative and phenomenological evidence shows that damage to the intercostobrachial nerve and axillary lymph node dissection often cause persistent sensory abnormalities, including numbness, stiffness, and tingling[34, 35]. When superimposed with adjuvant therapies like radiotherapy and chemotherapy, these physical discomforts serve as constant somatic reminders of "patient status" and "physical deficiency," thereby threatening body image integrity. To maintain family functioning or avoid being perceived as a "burden," patients frequently suppress their genuine distress regarding physical discomfort[36], falling into conflict over emotional expression and escalating alexithymia[37]. This process of internalizing somatic suffering and withdrawing socially ultimately translates into internalized shame regarding bodily incompleteness.Consequently, clinical management must extend beyond oncological endpoints to include early assessment and intervention for postoperative complications, particularly upper-limb functional rehabilitation. Clinicians should guide patients through standardized postoperative functional exercises, monitor upper-limb adverse outcomes, and mitigate the persistent psychological stress of physical dysfunction through active symptom management and health education[38]. Furthermore, evidence-based psychological strategies—such as Cognitive Behavioral Therapy (CBT), mindfulness-based interventions, and emotional disclosure exercises—should be integrated alongside peer support groups, family communication guidance, and body image restoration interventions (e.g., breast prosthesis fitting) within a multidisciplinary framework[37].

#### Psychological internal resources

Self-compassion, as a positive internal personal resource, helps individuals block the negative cycle of "self-criticism to emotional exhaustion" when dealing with physical defects and psychological trauma. The results of this study indicated that the level of self-compassion was a protective factor against stigma in post-mastectomy patients with breast cancer (*OR* = 0.874, *P*<0.001), which is consistent with the findings of Wang et al.[13]. Correlation analysis further revealed that the total SCS score was significantly and negatively correlated with the total SIS score and all its dimensions (*r*=-0.296∼-0.163, all *P*<0.05). Regarding positive dimensions, common humanity exhibited significant negative correlations with both the total SIS score and all its dimensions (*r*=-0.256∼-0.165, all *P*<0.05). Self-kindness was significantly and negatively correlated with the total SIS score (*r*=-0.169), financial insecurity (*r*=-0.254), and social rejection (*r*=-0.148) (all *P*<0.05), whereas its correlations with internalized shame (*r*=-0.061) and social isolation (*r*=-0.110) were not statistically significant (*P*>0.05). Mindfulness showed significant negative correlations with financial insecurity (*r*=-0.170) and social rejection (*r*=-0.134) (both *P*<0.05), but showed no statistically significant correlations with the total SIS score (*r*=-0.113), internalized shame (*r*=-0.001), or social isolation (*r*=-0.069) (all *P*>0.05). In terms of negative dimensions (self-judgment, over-identification, and isolation), each showed significant positive correlations with the total SIS score and all its dimensions (*r*=0.131∼0.260, all *P*<0.05), suggesting that unhelpful/negative self-cognitions aggravate the experience of stigma. Furthermore, Özönder Ünal et al.[39]reported that cancer patients scored higher on the negative dimensions of self-compassion (self-judgment, isolation, and over-identification) than on the positive dimensions (self-kindness, common humanity, and mindfulness). This psychological "negative bias" isolates patients, impairs their ability to form emotional connections, drives self-blame, and hinders peaceful acceptance of current experiences[40]. From a psychological perspective, post-mastectomy stigma functions as both a cognitive attitude and a psychological stress response shaped by life events, cognitive appraisal, coping mechanisms, and social support[41]. As an adaptive cognitive appraisal strategy, self-compassion directly regulates coping behaviors[40]. Evidence indicates that stigma arises when external stressors trigger perceived threats, activating the brain’s self-defense system. Conversely, self-compassion activates the affiliation system, stimulating the release of oxytocin and endorphins[42, 43]. This neurobiological cascade generates feelings of safety, warmth, and protection, thereby alleviating stigma.Consequently, clinicians should integrate self-compassion cultivation into routine clinical care. Beyond encouraging mindfulness and self-kindness, targeted interventions—such as Cognitive Behavioral Therapy (CBT) to restructure automatic negative thoughts, peer group sharing to demystify shame, self-acceptance writing exercises, and mindfulness awareness training—may establish a refined, stratified interventional pathway.

#### External social resources and cross-level resource interactions

Our results revealed that perceived social support was an independent protective factor against elevated stigma among post-mastectomy breast cancer patients (*OR*=0.914, *P*=0.003; *OR*=0.872, *P*=0.001). Total PSSS scores were negatively correlated with total SIS scores and all subscale scores. Notably, the absolute correlation coefficients with total SIS scores decreased progressively from "other support" (*r*=-0.297), to "family support" (*r*=-0.234), and "friends support" (*r*=-0.216). These findings align with He et al.[44] and the systematic review by Tang et al.[32], which emphasized social support as a critical buffer against cancer-related stigma. In this study, "other support" (derived from supervisors, colleagues, and broader social networks) exhibited a stronger inverse association with stigma than family or friend support. This suggests that societal recognition in workplace and community settings constitutes a pivotal external resource, echoing Su et al.[45], who confirmed that social validation upon returning to work significantly mitigates disease stigma, enhances social adaptation, and improves work behaviors[46]. From the perspective of Conservation of Resources (COR) theory, these findings underscore the vital role of external resource reservoirs in buffering resource depletion spirals. Consequently, clinical interventions must extend beyond individual psychological therapy to actively assess the integrity of patients’ social support networks. For patients with sparse social support, healthcare providers should mobilize family caregiving while connecting patients to peer support groups, community rehabilitation services, and financial aid policies to rebuild multidimensional social ties.

Contrary to our hypothesis grounded in the resource caravan model, the interaction between self-compassion and perceived social support was not statistically significant. This finding indicates that, within this cohort of post-mastectomy breast cancer patients, internal psychological resources and external environmental resources mitigate stigma through independent, parallel pathways rather than synergistic mechanisms. This lack of interaction likely stems from the distinct constructs evaluated by the two instruments: SCS captures intrapersonal emotional regulation, whereas the PSSS assesses external resource availability. These resources alleviate stigma through complementary mechanisms—self-compassion attenuates internalized shame and self-blame via cognitive reappraisal, while perceived social support buffers against social rejection and isolation through emotional and instrumental assistance. Because these pathways operate via distinct domains, they function independently without generating a multiplicative effect. Importantly, the absence of an interaction effect does not diminish the clinical value of either resource. Instead, it highlights that clinical interventions should simultaneously and independently foster self-compassion and strengthen social support networks, as each provides unique protective benefits against stigma.

#### Limitations

Several limitations of the present study warrant consideration. First, due to the single-center convenience sampling and cross-sectional design, the study cohort was recruited exclusively from tertiary hospitals in a specific region, reflecting psychological states at a single postoperative time point. This design inherently constrains the generalizability of the findings and limits our capacity to infer longitudinal causal dynamics among variables. Future multi-center, large-sample prospective studies are encouraged to evaluate the stability and generalizability of the identified latent profiles. Second, data collection relied primarily on self-report instruments. Although statistical evaluation revealed no threat of severe common method bias, self-reported responses remain susceptible to social desirability bias and memory-related cognitive distortion. Future research could incorporate objective physiological markers or qualitative interviews to achieve a more comprehensive assessment. Finally, the interaction between self-compassion and perceived social support failed to reach statistical significance, suggesting either the absence of a robust synergistic mechanism or insufficient statistical power to detect subtle interaction effects given the current sample size. Future studies employing larger sample sizes, longitudinal tracking, or structural equation modeling (SEM) are needed to further elucidate the complex, dynamic interplay between internal and external resources in shaping cancer-related stigma.

## Conclusions

Grounded in Conservation of Resources (COR) theory, this study applied latent profile analysis to demonstrate significant latent heterogeneity in stigma among post-mastectomy breast cancer patients, identifying three distinct subgroups: the low-stigma profile, moderate-stigma profile, and high-stigma profile. Our findings confirm that age, educational level, employment status, monthly income, affected upper-limb paresthesia, and clinical TNM stage are key predictors of stigma profile membership. Crucially, higher levels of self-compassion and perceived social support act as critical protective buffers that mitigate the risk of high stigma. These results underscore the imperative for precision, stratified nursing interventions tailored to distinct psychological phenotypes. Healthcare providers should focus clinical vigilance on high-risk patient cohorts, actively implementing integrated psychological strategies that cultivate self-compassion and bolster external social support networks. Such targeted approaches are essential for alleviating disease-related stigma, facilitating psychological adaptation, and fostering long-term social function reconstruction following modified radical mastectomy.

## Data Availability

All relevant data are within the paper and its Supporting Information files.

## Acknowledgments

We extend our thanks to the medical and nursing staff at Deyang City People’s Hospital Breast Surgery Department for their assistance with participant recruitment and data collection. We also thank Hospital statistician for providing statistical advice during data analysis, as well as all the patients who voluntarily participated in this study.

## Supporting information

**S1 Fig. Ethical Approval Document.**

**S1 Table. Analysis Data Table of Influencing Factors.**

**S2 Table. Potential category classification.**

**S1 File. Mplus latent profile.**

## References

1. Sung H, Filho AM, Laversanne M, Ferlay J, Siegel RL, Soerjomataram I, et al. Global cancer statistics 2024: GLOBOCAN estimates of incidence and mortality worldwide for 34 cancers in 186 countries. CA Cancer J Clin. 2026;76(4):e70090. 10.3322/caac.70090. PMID: 42417444.

2. Zheng R, Sun K, Han B, Li L, Chen R, Wang S, et al. Cancer incidence and mortality in China, 2024. Journal of the National Cancer Center. 2026. 10.1016/j.jncc.2025.10.003.

3. LaRiviere MJ, Chao HH, Doucette A, Kegelman TP, Taunk NK, Freedman GM, et al. Factors Associated With Fatigue in Patients with Breast Cancer Undergoing External Beam Radiation Therapy. Pract Radiat Oncol. 2020;10(6):409–22. Epub 20200609. 10.1016/j.prro.2020.05.011. PMID: 32531443.

4. Bean HR, Diggens J, Ftanou M, Weihs KL, Stanton AL, Wiley JF. Insomnia and Fatigue Symptom Trajectories in Breast Cancer: A Longitudinal Cohort Study. Behavioral Sleep Medicine. 2021;19(6):814–27. 10.1080/15402002.2020.1869005. PMID: 33470847.

5. Liu YH, Liu ZZ, Wang X, Wang GQ, Yu ZG. Expert consensus and surgical operation guidelines for breast cancer modified radical mastectomy (2018 edition) . Chinese Journal of Practical Surgery. 2018;38(8):851–854. 10.19538/j.cjps.issn1005-2208.2018.08.03.

6. Suwankhong D, Liamputtong P. Breast Cancer Treatment: Experiences of Changes and Social Stigma Among Thai Women in Southern Thailand. Cancer Nurs. 2016;39(3):213–20. 10.1097/ncc.0000000000000255. PMID: 25881809.

7. Wu J, Zeng N, Wang L, Yao L. The stigma in patients with breast cancer: A concept analysis. Asia-Pacific Journal of Oncology Nursing. 2023;10(10):100293. 10.1016/j.apjon.2023.100293. PMID: 37886719.

8. Chu Q, Wong CCY, Chen L, Shin LJ, Chen L, Lu Q. Self-stigma and quality of life among Chinese American breast cancer survivors: A serial multiple mediation model. Psychooncology. 2021;30(3):392–9. Epub 20201127. 10.1002/pon.5590. PMID: 33175446.

9. Fife BL, Wright ER. The Dimensionality of Stigma: A Comparison of Its Impact on the Self of Persons with HIV/AIDS and Cancer. Journal of Health and Social Behavior. 2000;41(1):50–67. doi: 10.2307/2676360. PMID: 10750322.

10. Burić I, Wang H. Relationships among teacher enjoyment, emotional labor, and perceived student engagement: A daily diary approach. Journal of School Psychology. 2024;103:101271. 10.1016/j.jsp.2023.101271. PMID: 38432728

11. Hobfoll SE, Halbesleben J, Neveu JP, Westman M. Conservation of Resources in the Organizational Context: The Reality of Resources and Their Consequences. Annual Review of Organizational Psychology and Organizational Behavior. 2018;5(1):103–128. 10.1146/annurev-orgpsych-032117-104640.

12. Neff, KD. Self-Compassion: An Alternative Conceptualization of a Healthy Attitude Toward Oneself. Self and Identity. 2003;2(2):85–101. 10.1080/15298860309032.

13. Wang JH, Bai P, Wei YT. The Status of Postoperative Stigma of Breast Cancer Patients and Its Correlation with Self-compassion in a Grade-A Tertiary Hospital in Beijing. Medicine and Society. 2024;37(6):138–144. 10.13723/j.yxysh.2024.06.020.

14. Liu CL, Qin WZ, Xu LZ, Zhang J, Gao ZR, et al. The mediating effect of depression between the perception of social support and life satisfaction among the elderly in Tai’an city. Modern Preventive Medicine. 2022;49(3):466–471.

15. Yu L, Fan HF, Li XL, Du J, Jia ZH. Current status and influencing factors of stigma in postoperative patients with head and neck cancer. Chinese Nursing Research. 2024;38(5):915–920. 10.12102/j.issn.1009-6493.2024.05.029

16. Yin K, Peng J, Zhang J. The application of latent profile analysis in organizational behavior research. Advances in Psychological Science. 2020;28(7):1056–70. http://dx.chinadoi.cn/10.3724/SP.J.1042.2020.01056.

17. Li S, Wang X, Wang M, Jiang Y, Mai Q, Wu J, et al. Association between stigma and sleep quality in patients with breast cancer: A latent profile and mediation analysis. Eur J Oncol Nurs. 2023;67:102453. Epub 20231027. doi: 10.1016/j.ejon.2023.102453. PubMed PMID: 37951070.

18. Breast Cancer Professional Committee of China Anti-Cancer Association; Breast Oncology Group of Chinese Society of Clinical Oncology, Chinese Medical Association. Guidelines and standards for the diagnosis and treatment of breast cancer by the China Anti-Cancer Association (2026 edition). China Oncology. 2025;35(12):1157–1255. http://dx.chinadoi.cn/10.19401/j.cnki.1007-3639.2025.12.009.

19. Tein JY, Coxe S, Cham H. Statistical Power to Detect the Correct Number of Classes in Latent Profile Analysis. Struct Equ Modeling. 2013;20(4):640–57. 10.1080/10705511.2013.824781. PMID: 24489457.

20. Lewis SC. Sample size calculations in clinical research. Taylor & Francis; 2009. 10.1080/02664760802366775

21. Pan AW, Chung LI, Fife BL, Hsiung PC. Evaluation of the psychometrics of the Social Impact Scale: a measure of stigmatization. Int J Rehabil Res.2007;30(3):235. 10.1097/MRR.0b013e32829fb3db. PMID: 17762770.

22. Chen J, Yan LS, Zhou LH. Reliability and Validity of Chinese Version of Self-compassion Scale. Chinese Journal of Clinical Psychology. 2011;19(6):734–736. 10.16128/j.cnki.1005-3611.2011.06.006.

23. Zimet GD, Powell SS, Farley GK, Werkman S, Berkoff KA. Psychometric characteristics of the Multidimensional Scale of Perceived Social Support. J Pers Assess. 1990;55(3-4):610–7. 10.1080/00223891.1990.9674095. PMID: 2280326.

24. Jiang QJ. Perceived Social Support Scale. Chinese Journal of Behavioral Medical Science. 2001;10:41–43.

25. Mai WJ. Analysis of influencing factors of stigma among postoperative breast cancer patients. Henan Journal of Surgery. 2025;31(6):102–104. 10.16193/j.cnki.hnwk.2025.06.049.

26. Jiang MF, Gao J, Yang L, Lu YM, Qian J, Li JZ. Mediating effect of self-disclosure between stigma and loneliness in breast cancer survivors. 2023;48(4):516–20. 10.13898/j.cnki.issn.1000-2200.2023.04.023.

27. Li SS. Influencing factors and risk predictionmodel construction of high stigma inpatients with breast cancer after modifiedradical mastectomy. Chinese Journal of Social Medicine. 2026;38(10):32–35+39. http://dx.chinadoi.cn/10.3969/j.issn.1672-0369.2026.10.009.

28. Yin CL, Qu HL, Yang FG, Ren HQ, Leng HP. Psychosocial adjustment and influencing factors of young patients with breast cancer. Journal of Nursing Administration. 2019;19(7):457–461. http://dx.chinadoi.cn/10.3969/j.issn.1671-315x.2019.07.001.

29. Yang R, Yan R, Yuan F, Liu DL, Wang WM, Qu YX. The present situation of stigma and its influencing factors in ovarian cancer patients. Journal of Nurses Training. 2020;35(23):2128–2132. http://dx.chinadoi.cn/10.16821/j.cnki.hsjx.2020.23.004.

30. Qiu JJ, Li P, Huang LJ, Tao J. Correlation Study on Social Restrictions and Stigma in Young Breast Cancer Patients. Medicine and Philosophy. 2023;44(18):47–51. http://dx.chinadoi.cn/10.12014/j.issn.1002-0772.2023.18.11.

31. Wang Y, Zhu ZH, Zhang RX, et al. Mediating effect of social anxiety and stigma on family caring and loneliness in breast cancer patients. Chinese Nursing Research. 2024;38(4):571–576. http://dx.chinadoi.cn/10.12102/j.issn.1009-6493.2024.04.002.

32. Tang WZ, Yusuf A, Jia K, Iskandar YHP, Mangantig E, Mo XS, et al. Correlates of stigma for patients with breast cancer: a systematic review and meta-analysis. Support Care Cancer. 2022;31(1):55. Epub 20221217. 10.1007/s00520-022-07506-4. PMID: 36526859.

33. Tan C, Zhong C, Mei R, Yang R, Wang D, Deng X, et al. Stigma and related influencing factors in postoperative oral cancer patients in China: a cross-sectional study. Support Care Cancer. 2022;30(6):5449–58. Epub 20220319. 10.1007/s00520-022-06962-2. PMID: 35305161.

34. Koçan S, Gürsoy A. Body Image of Women with Breast Cancer After Mastectomy: A Qualitative Research. J Breast Health. 2016;12(4):145–50. Epub 20161001. 10.5152/tjbh.2016.2913. PMID: 28331752.

35. Everaars KE, Welbie M, Hummelink S, Tjin EPM, de Laat EH, Ulrich DJO. The impact of scars on health-related quality of life after breast surgery: a qualitative exploration. J Cancer Surviv. 2021;15(2):224–33. Epub 20200820. 10.1007/s11764-020-00926-3. PMID: 32816201.

36. Rao X, Wang HX, Shi BZ, Zhang J. Conflict over emotional expression in breast cancer patients based on the ABC-X model: A mixed-methods study on current status and influencing factors. Journal of Nursing Science. 2026;41(4):85–90. 10.3870/j.issn.1001-4152.2026.04.085.

37. Liu Y, Du Q, Jiang Y. Prevalence of alexithymia in cancer patients: a systematic review and meta-analysis. Support Care Cancer. 2023;31(12):675. Epub 20231107. 10.1007/s00520-023-08106-6. PMID: 37932546.

38. Wang YF, Wang YL, Yang GW, Yuan Y, Li Q, Kong P. Current situation and influencing factors ofself-management activation amongpatients with Breast Cancer-RelatedLymphedema. Chinese Nursing Management. 2024;24(9):1329– 1334.

39. Ozonder Unal I, Ordu C. Alexithymia, Self-Compassion, Emotional Resilience, and Cognitive Emotion Regulation: Charting the Emotional Journey of Cancer Patients. Current Oncology. 2023; 30(10): 8872–87. 10.3390/curroncol30100641.

40. Neff KD. Self-Compassion, Self-Esteem, and Well-Being. Social & Personality Psychology Compass. 2011.

41. Zheng J, Yang LP, Ma HZ, Zhu JH, Li G. Research progress on correlation betweenchildren with emotion and cognitiveimpairment and their mother’spsychological stress during pregnancy. China Journal of Traditional Chinese Medicine and Pharmacy.

42. Gilbert P. Compassion : Conceptualisations, Research and Use in Psychotherapy. 2005.

43. Kerstin Us-M, Linda H, Maria P. Self-soothing behaviors with particular reference to oxytocin release induced by non-noxious sensory stimulation. Frontiers in Psychology. 2014;5:1529. 10.3389/fpsyg.2014.01529. PMID: 25628581.

44. He J, Li N, Shi YJ. The mediating role of disease stigma in the appreciationof young breast cancer patients regarding the relationshipbetween social support and social alienation. Health Research. 2023;43(6):655–9. http://dx.chinadoi.cn/10.19890/j.cnki.issn1674-6449.2023.06.011.

45. Su XQ, Wu TT, Zhang HY, Shi Q, Xu Y, Kuai BX, et al. Mediating and moderating factors between stigma and adaptability to return to work for cancer survivors. Sci Rep. 2024;14(1):30944. Epub 20241228. 10.1038/s41598-024-82013-6. PubMed PMID: 39730772.

46. Zhang YD, Liu RH, Han F. The impact of stigma on job behavior among breast cancer survivors after return to work: a cross-sectional study. Journal of Nursing Science. 2020;35(3):67–70. http://dx.chinadoi.cn/10.3870/j.issn.1001-4152.2020.03.067.

